# A cross-sectional study of the relationship between exercise, physical activity, and health-related quality of life among Japanese workers during the COVID-19 pandemic

**DOI:** 10.1101/2021.06.15.21259002

**Authors:** Ryosuke Sugano, Kazunori Ikegami, Hisashi Eguchi, Mayumi Tsuji, Seiichiro Tateishi, Tomohisa Nagata, Shinya Matsuda, Yoshihisa Fujino, Akira Ogami, for the CORoNaWork Project

## Abstract

**Background:** Exercise and physical activity positively affect physical and mental health, and healthy workers contribute to increased work productivity. This study aimed to investigate time spent on exercise during leisure time and physical activity, including time at work, in relation to health-related quality of life (HRQOL) among Japanese workers.

**Methods:** An Internet-based national health survey –Collaborative Online Research on Novel-coronavirus and Work study (CoRoNaWork study) – was conducted among 33,087 Japanese workers in December 2020. After excluding invalid responses, 27,036 participants were categorized into four and five groups according to exercise time and physical activity time, respectively. Each group’s scores were compared on each of the four questions on the Japanese version of the Centers for Disease Control and Prevention Health-Related Quality of Life (CDC HRQOL-4) between each group using a linear mixed model. Age-sex adjusted and multivariate models were used to compare each index of the CDC HRQOL-4.

**Results:** The more time spent exercising, the better the self-rated health. Self-rated health was better and unhealthy days were fewer in the group that exercised more than a certain amount of time than in the group that spent almost no time on exercising. As for physical activity, the group that spent more than 120 minutes/day or that almost never engaged in physical activity had lower self-rated health. The group with more than 120 minutes/day of physical activity also had more unhealthy days.

**Conclusions:** The results of the present study suggest that exercise habits improve workers’ HRQOL. Interventions to encourage daily exercise even for a short period of time may be an effective strategy to improve workers’ health and productivity.

## Background

People who frequently engage in physical activity and have good exercise habits are known to have lower morbidity and mortality rates for all-cause mortality, ischemic heart disease, hypertension, diabetes, obesity, osteoporosis, and colon cancer, as well as improved depression and anxiety symptoms and reduced prevalence of panic attacks, social phobia, and plaque phobia [1-4]. However, about one in four men and one in three women worldwide do not engage in the physical activity and exercise, necessary to maintain good health, as recommended by the World Health Organization (WHO) [5]. Surveys conducted in Japan reported that 32% of men and 26% of women exercised for 30 minutes or more at least twice a week [6], and that 33% of workers participated in regular exercise [7].

Some studies show that absenteeism due to health problems decreased with increased physical activity [8], that regular aerobic exercise and strength training prevented work productivity loss due to presenteeism [9], and that increased physical activity reduced work restrictions [10]. These findings suggest that exercise and physical activity have a positive impact on work productivity. Thus, workers need to have good exercise habits, not only for their health but also for their work performance.

The global epidemic of the coronavirus disease 2019 (COVID-19) will continue to significantly impact daily life, work, and healthcare worldwide in 2021. Studies have shown that public health restrictions related to COVID-19 have reduced the amount of time spent outdoors [11], and that 70% of people surveyed showed a moderate or greater change in physical activity during the period when voluntary restraint was recommended [12]. In Japan, the government declared a state of emergency in April 2020, and people were asked to refrain from going out. In addition, to prevent the spread of infection, facilities where many people gather, such as department stores, movie theaters, gyms, and sports grounds, were requested to close [13]. These measures affected people’s exercise habits and reduced the time spent in physical activities, such as walking.

In the present study, we focused on the relationship between physical activity and health-related quality of life (HRQOL). This multidimensional and broad concept includes health, work, culture, and religion, and is limited to physical and mental health factors. The Centers for Disease Control and Prevention (CDC) defined HRQOL as an individual’s or group’s perceived physical and mental health over time [14]. Four core components have been used to assess HRQOL: self-rated health, the number of days in the past 30 days that a person was physically or mentally unhealthy, and the number of days when unhealthy conditions limited a person’s activities. The CDC HRQOL-4, developed by the CDC as a tool for public health surveillance, measures these four factors [15]. International studies have provided support for the reliability and validity of the CDC HRQOL-4. In 2020, the Japanese version of the CDC HRQOL-4 was developed, and evaluated for its reliability and validity for use with workers [16]. Furthermore, because the CDC HRQOL-4 was found to be associated with indicators of work functioning impairment [17], the measurement of HRQOL is helpful for both health and labor-management assessment of workers.

Understanding the relationship between time spent exercising during leisure time, time spent engaged in physical activity including work, and HRQOL will provide important information for developing interventions to improve workers’ health and work productivity. Furthermore, we thought that it would be useful to investigate these relationships during the COVID-19 pandemic, mainly because physical activity tends to be restricted. As such, this study investigated the relationship between exercise, physical activity, and HRQOL using a large-scale Internet survey of workers conducted during the COVID-19 pandemic.

## Methods

### Study design and setting

The present study was cross-sectional and used a part of the baseline data of a prospective cohort study that was conducted by a research group at the University of Occupational and Environmental Health in Japan. The study is called the Collaborative Online Research on Novel-coronavirus and Work study (CORoNaWork study). A Japanese Internet survey company—Cross Marketing Inc.

Tokyo—implemented the study, which involved a self-administered questionnaire survey. Baseline survey data was collected from December 22–25, 2020. Incidentally, the baseline survey occurred during the third wave of the pandemic when the number of COVID-19 infections and deaths was overwhelmingly higher than in the first and second waves; therefore, Japan was on high alert. The details of the study protocol are provided by Fujino et al. [18].

### Participants

The participants were between 20 and 65 years and working at the time of the baseline survey. A total of 33,087 adults participated in the CORoNaWork study, which employed stratified cluster sampling by gender, age, region, and occupation. A database of 27,036 individuals was created by excluding 6,051 who were determined to have provided invalid responses.

### Questionnaire

The questionnaire items are described in detail by Fujino et al. [18]. We used questionnaire data on sex, age, presence of illness under medical treatment, average exercise time during leisure time per day, average physical activity time including work/day, educational background, area of participants’ residence, job type, working hours per day, telecommuting frequency, and HRQOL. The CDC HRQOL-4 was used to assess overall HRQOL, including the following four items: (a) self-rated health, (b) the number of physically unhealthy days in the past 30 days, (c) the number of mentally unhealthy days in the past 30 days, and (d) the number of days with activity limitation in the past 30 days. Self-rated health was recorded as excellent, very good, good, fair, or poor [19]. We used the Japanese version of the CDC HRQOL-4 developed by Japanese epidemiologists and occupational physicians [17].

### Variables

#### Outcome variables

The outcome variables were CDC HRQOL-4 scores for self-rated health (5-point Likert scale: 1 = excellent, 2 = very good, 3 = good, 4 = fair, and 5 = poor) and the number of days in the past 30 days reported for physically unhealthy days, mentally unhealthy days, and activity limitation days.

#### Predictor variables

We classified the participants into four groups according to their exercise time during leisure time: ≥60 minutes/day, 30–59 minutes/day, 1–29 minutes/day, and almost never. Participants were also classified into five groups according to their physical activity time, including work: ≥120 minutes/day, 60–119 minutes/day, 30–59 minutes/day, 1–29 minutes/day, and almost never. These variables were used as the predictor variables.

#### Potential confounding variables

The following items were used as confounding factors. Sex, age (20–29, 30–39, 40–49, 50–59, and ≥ 60 years of age), educational background (junior or senior high school, junior college or vocational school, university or graduate school), and presence of illness under medical treatment were used as personal characteristics. Occupation (regular employee, manager, executive, public service worker, temporary worker, freelancer or professional, other), telecommuting frequency (4 days/week or more, 2–3 days/week, 1 day/week or less, almost never), working hours per day (less than 8 hours/day, 8–9 hours/day, 9–11 hours/day, 11 hours/day or more) were used as work-related factors. The prefecture of the participants’ residence was also used.

### Statistical method

We used a linear mixed model (LMM) to analyze the relationships between the four items of the CDC HRQOL-4 and the four groups of exercise time, and the five groups of physical activity time. Two models were analyzed for each predictor variable. In the age-sex adjusted model, we treated the four classifications of exercise time, the five classifications of physical activity time, age, and sex as fixed effects, and treated the prefecture of residence as random effects. In the multivariate model, we added personal characteristics and work-related variables to the fixed effects of the age-sex adjusted model. The Bonferroni test was used for post-hoc testing after LMM. The estimated marginal means (EMM) of the four items of the CDC HRQOL-4 by the four groups of exercise time and the five groups of physical activity time were calculated by adjusting for the dependent variable in each model. In all the tests, the threshold for significance was set at P < 0.05. We used SPSS 25.0 J analytical software (IBM, NY, USA) for statistical analyses.

## Results

### Participants and descriptive data

As for exercise during leisure time, 3,202 (11.8%) participants spent 60 minutes/day or more, 4,210 (15.6%) spent 30–59 minutes/day, 6,123 (22.6%) spent 1–29 minutes/day, and 13,501 (49.9%) reported they almost never exercised. As for physical activity time including work, 6,843 (25.3%) participants spent more than 120 minutes/day, 3,062 (11.3%) spent 60–119 minutes/day, 4,257 (15.7%) spent 30–59 minutes/day, 4,770 (17.6%) spent 1–29 minutes/day, and 8,104 (30.0%) reported they almost never spent time engaged in physical activity (Table 1).

**Table 1.** Participants’ characteristics.

| Items | n | (%) |
| --- | --- | --- |
| N | 27036 | (100) |
| Sex, male | 13814 | (51.1) |
| Age |  |  |
| 20–29 | 1905 | (7.0) |
| 30–39 | 4858 | (18.0) |
| 40–49 | 8011 | (29.6) |
| 50–59 | 9012 | (33.3) |
| ≥60 | 3250 | (12.0) |
| Education |  |  |
| Junior or senior high school | 7321 | (27.1) |
| Junior college or vocational school | 6544 | (24.2) |
| University or graduate school | 13171 | (48.7) |
| Presence of illnesses that require hospital treatment. | 9510 | (35.2) |
| Job type |  |  |
| Desk work | 13468 | (49.8) |
| Hospitality work | 6927 | (25.6) |
| Manual work | 6641 | (24.6) |
| Telecommuting frequency |  |  |
| ≥4 days/week | 2790 | (10.3) |
| 2–3 days/week | 1477 | (5.5) |
| ≤1 day/week | 1493 | (5.5) |
| None | 21276 | (78.7) |
| Working hours per day |  |  |
| <8 hours/day | 5334 | (19.7) |
| 8–9 hours/day | 14848 | (54.9) |
| 9–11 hours/day | 5541 | (20.5) |
| ≥11 hours/day | 1313 | (4.9) |
| Exercise time in leisure time |  |  |
| ≥60 minutes/day | 3202 | (11.8) |
| 30–59 minutes/day | 4210 | (15.6) |
| 1–29 minutes/day | 6123 | (22.6) |
| Almost never | 13501 | (49.9) |
| Physical activity time including work |  |  |
| ≥120 minutes/day | 6843 | (25.3) |
| 60–119 minutes/day | 3062 | (11.3) |
| 30–59 minutes/day | 4257 | (15.7) |
| 1–29 minutes/day | 4770 | (17.6) |
| Almost never | 8104 | (30.0) |

### Comparison of the scores on the CDC HRQOL-4 among the exercise time groups

In the age-sex adjusted and multivariate models, we used LMM to compare the CDC-HRQOL-4 scores for each of the four groups of exercise time (Tables 2 and 3). The two models showed a main effect for self-rated health. Post hoc tests showed that shorter exercise time tended to be associated with significantly lower self-rated health (P = 0.016 for the pair of 30–59 minutes/day group and 1–29 minutes/day group in the age-sex adjusted model, and P < 0.001 for the others; P = 0.017 for the pair of 30–59 minutes/day group and 1–29 minutes/day group in the multivariate model, and P < 0.001 for the others). The two models also showed the main effect for the three items on physically unhealthy days, mentally unhealthy days, and days with activity limitation. Post hoc tests showed that the almost never group had significantly more physically unhealthy days than the ≥ 60 minutes/day group and the 30–59 minutes/day group (P = 0.002 and P < 0.001 in the age-sex adjusted model, respectively, and P = 0.008 and P < 0.001 in the multivariate model, respectively), and significantly more mentally unhealthy days than the other three groups (P < 0.001 for both the age-sex adjusted model and multivariate model). The almost never group had significantly more days with activity limitation than the 30–59 minutes/day group and the 1–29 minutes/day group (P < 0.001 and P = 0.028 in the age-sex adjusted model, and P = 0.001 and P = 0.047 in the multivariate model, respectively).

**Table 2.** Exercise time in leisur**e time and physical activity time including work**

| Parameters | Items/Options | Age-sex adjusted |  |  |  | Multivariate* |  |  |  |
| --- | --- | --- | --- | --- | --- | --- | --- | --- | --- |
|  |  | EMM | 95% CI | p | P for interaction | EMM | 95% CI | p | P for interaction |
| Self-rated health | Exercise time in leisure time |  |  |  |  |  |  |  |  |
|  | ≥60 minutes/day | 3.25 | [3.22-3.29] | <0.001 |  | 3.34 | [3.30-3.38] | <0.001 |  |
|  | 30–59 minutes/day | 3.35 | [3.32-3.38] |  |  | 3.43 | [3.40-3.47] |  |  |
|  | 1–29 minutes/day | 3.41 | [3.39-3.44] |  |  | 3.49 | [3.46-3.53] |  |  |
|  | Almost never | 3.55 | [3.52-3.57] |  |  | 3.62 | [3.59-3.65] |  |  |
|  | Physical activity time including work |  |  |  | 0.044 |  |  |  | 0.059 |
|  | ≥120 minutes/day | 3.42 | [3.39-3.44] | <0.001 |  | 3.48 | [3.45-3.51] | <0.001 |  |
|  | 60–119 minutes/day | 3.33 | [3.30-3.37] |  |  | 3.42 | [3.38-3.45] |  |  |
|  | 30–59 minutes/day | 3.35 | [3.32-3.38] |  |  | 3.43 | [3.40-3.47] |  |  |
|  | 1–29 minutes/day | 3.38 | [3.35-3.42] |  |  | 3.47 | [3.43-3.51] |  |  |
|  | Almost never | 3.47 | [3.44-3.51] |  |  | 3.56 | [3.51-3.60] |  |  |

EMM: estimated marginal mean, CI: confidence interval.
\*Multivariate are adjusted for age, sex, education, job type, telecommuting frequency, and working hours per day.

**Table 3.**
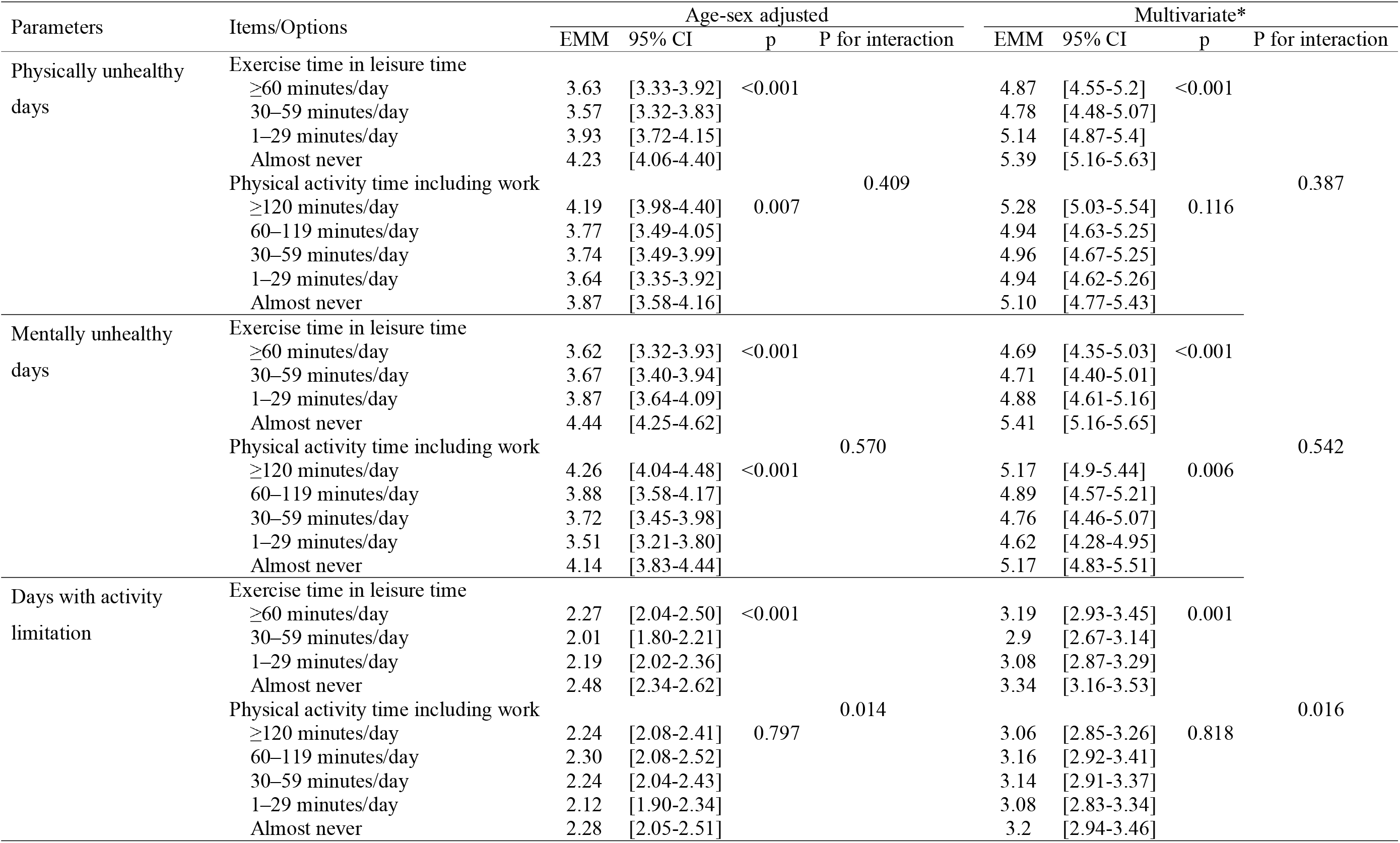
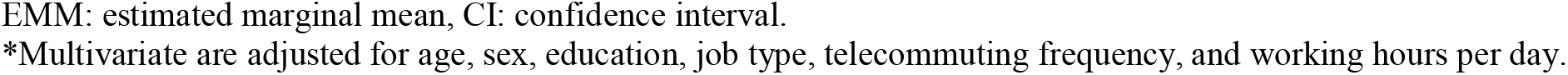
Unhealthy days, exercise time in leisure time, and physical activity time including work.

| Parameters | Items/Options | Age-sex adjusted |  |  |  | Multivariate* |  |  |  |
| --- | --- | --- | --- | --- | --- | --- | --- | --- | --- |
|  |  | EMM | 95% CI | p | P for interaction | EMM | 95% CI | p | P for interaction |
| Physically unhealthy days | Exercise time in leisure time |  |  |  |  |  |  |  |  |
|  | ≥60 minutes/day | 3.63 | [3.33-3.92] | <0.001 |  | 4.87 | [4.55-5.2] | <0.001 |  |
|  | 30–59 minutes/day | 3.57 | [3.32-3.83] |  |  | 4.78 | [4.48-5.07] |  |  |
|  | 1–29 minutes/day | 3.93 | [3.72-4.15] |  |  | 5.14 | [4.87-5.4] |  |  |
|  | Almost never | 4.23 | [4.06-4.40] |  |  | 5.39 | [5.16-5.63] |  |  |
|  | Physical activity time including work |  |  |  | 0.409 |  |  |  | 0.387 |
|  | ≥120 minutes/day | 4.19 | [3.98-4.40] | 0.007 |  | 5.28 | [5.03-5.54] | 0.116 |  |
|  | 60–119 minutes/day | 3.77 | [3.49-4.05] |  |  | 4.94 | [4.63-5.25] |  |  |
|  | 30–59 minutes/day | 3.74 | [3.49-3.99] |  |  | 4.96 | [4.67-5.25] |  |  |
|  | 1–29 minutes/day | 3.64 | [3.35-3.92] |  |  | 4.94 | [4.62-5.26] |  |  |
|  | Almost never | 3.87 | [3.58-4.16] |  |  | 5.10 | [4.77-5.43] |  |  |
| Mentally unhealthy days | Exercise time in leisure time |  |  |  |  |  |  |  |  |
|  | ≥60 minutes/day | 3.62 | [3.32-3.93] | <0.001 |  | 4.69 | [4.35-5.03] | <0.001 |  |
|  | 30–59 minutes/day | 3.67 | [3.40-3.94] |  |  | 4.71 | [4.40-5.01] |  |  |
|  | 1–29 minutes/day | 3.87 | [3.64-4.09] |  |  | 4.88 | [4.61-5.16] |  |  |
|  | Almost never | 4.44 | [4.25-4.62] |  |  | 5.41 | [5.16-5.65] |  |  |
|  | Physical activity time including work |  |  |  | 0.570 |  |  |  | 0.542 |
|  | ≥120 minutes/day | 4.26 | [4.04-4.48] | <0.001 |  | 5.17 | [4.9-5.44] | 0.006 |  |
|  | 60–119 minutes/day | 3.88 | [3.58-4.17] |  |  | 4.89 | [4.57-5.21] |  |  |
|  | 30–59 minutes/day | 3.72 | [3.45-3.98] |  |  | 4.76 | [4.46-5.07] |  |  |
|  | 1–29 minutes/day | 3.51 | [3.21-3.80] |  |  | 4.62 | [4.28-4.95] |  |  |
|  | Almost never | 4.14 | [3.83-4.44] |  |  | 5.17 | [4.83-5.51] |  |  |
| Days with activity limitation | Exercise time in leisure time |  |  |  |  |  |  |  |  |
|  | ≥60 minutes/day | 2.27 | [2.04-2.50] | <0.001 |  | 3.19 | [2.93-3.45] | 0.001 |  |
|  | 30–59 minutes/day | 2.01 | [1.80-2.21] |  |  | 2.9 | [2.67-3.14] |  |  |
|  | 1–29 minutes/day | 2.19 | [2.02-2.36] |  |  | 3.08 | [2.87-3.29] |  |  |
|  | Almost never | 2.48 | [2.34-2.62] |  |  | 3.34 | [3.16-3.53] |  |  |
|  | Physical activity time including work |  |  |  | 0.014 |  |  |  | 0.016 |
|  | ≥120 minutes/day | 2.24 | [2.08-2.41] | 0.797 |  | 3.06 | [2.85-3.26] | 0.818 |  |
|  | 60–119 minutes/day | 2.30 | [2.08-2.52] |  |  | 3.16 | [2.92-3.41] |  |  |
|  | 30–59 minutes/day | 2.24 | [2.04-2.43] |  |  | 3.14 | [2.91-3.37] |  |  |
|  | 1–29 minutes/day | 2.12 | [1.90-2.34] |  |  | 3.08 | [2.83-3.34] |  |  |
|  | Almost never | 2.28 | [2.05-2.51] |  |  | 3.2 | [2.94-3.46] |  |  |

EMM: estimated marginal mean, CI: confidence interval.
\*Multivariate are adjusted for age, sex, education, job type, telecommuting frequency, and working hours per day.

### Comparison of the scores on the CDC HRQOL-4 among the physical activity time groups

The age-sex adjusted model and multivariate model were also used to compare the CDC-HRQOL-4 scores for each of the five groups of physical activity time (Tables 2 and 3). The two models showed a main effect for self-rated health and mentally unhealthy days. In the age-sex adjusted model, the ≥ 120 minutes/day group had significantly lower self-rated health than the 60–119 minutes/day and 30–59 minutes/day groups (P = 0.001 and P = 0.012, respectively). Self-rated health was significantly lower in the almost never group than in the 60–119 minutes/day, 30–59 minutes/day, and 1–29 minutes/day groups (P < 0.001, P < 0.001, and P = 0.003, respectively). The ≥ 120 minutes/day group had significantly more mentally unhealthy days than the 30–59 minutes/day and 1–29 minutes/day groups (P = 0.010 and P < 0.001, respectively). The mentally unhealthy days were significantly more in the almost never group than in the 1–29 minutes/day group (P = 0.024). In the multivariate model, the almost never group had significantly lower self-rated health than the ≥ 120 minutes/day, 60–119 minutes/day, 30–59 minutes/day, and 1–29 minutes/day groups (P = 0.006, P < 0.001, P < 0.001, and P = 0.009, respectively), and the ≥ 120 minutes/day group had significantly more mentally unhealthy days than the 1–29 minutes/day group (P = 0.017). For physically unhealthy days, the age-sex adjusted model showed a main effect, and post hoc tests showed that the ≥ 120 minutes/day group had significantly more unhealthy days than the 1–29 minutes/day group (P = 0.015). No main effect was observed for days with activity limitation.

### Interaction of the exercise time groups and physical activity time groups

There was an interaction between exercise time and physical activity time in the two models for days with activity limitation. In the age-sex adjusted model, among the almost never group for time spent on exercise, those who almost never engaged in physical activity had significantly more days with activity limitation than the 30–59 minutes/day group (P = 0.005). In the multivariate model, among the almost never group for time spent on exercise, those who almost never engaged in physical activity had significantly more days with activity limitation than the 30–59 minutes/day group and 1–29 minutes/day group (P = 0.007, P = 0.046, respectively).

## Discussion

This study clarified the relationship between HRQOL (self-rated health and unhealthy days), exercise time during leisure time, and physical activity time including work. The relationship between exercise time and HRQOL showed that higher self-rated health was associated with increases in time spent exercising, even after adjusting for personal characteristics and work-related factors. In addition, the results suggest that an exercise habit of 30 minutes/day or more may help reduce the number of physically unhealthy days. Exercise, even for a short time every day, may effectively reduce mentally unhealthy days. The results of this study were similar to those of previous studies conducted on adults in various regions [20-23]. Another study has shown that moderate-to-vigorous physical activity of fewer than 30 minutes/day is associated with a reduced mortality rate [24], thereby implying that even short exercise sessions may contribute to better health and fewer mentally unhealthy days. These results provide valuable information while considering interventions to address exercise habits among workers.

As for the relationship between physical activity, including work, and HRQOL, no time spent or excessive time spent in physical activity was associated with lower self-rated health. In addition, the number of unhealthy days increased with the amount of time spent engaged in physical activity. The no time spent in physical activity result in part may be due to the work-related tasks of the individuals in the almost never group. In other words, it may be that more people in this group perform most of their work tasks in a sitting position. A study found a negative relationship between sitting time and self-rated health, although the sample was older adults [25]. It is also possible that the limited amount of time spent engaged in physical activity is due to a low sense of subjective health.

With regard to physical activity time and mental health, a previous study found that more exercise time during leisure time was negatively associated with depression but not significantly associated with more work-related physical activity time [26]. Furthermore, some studies have shown that the lower the socioeconomic status, the higher the work-related physical activity [27], and that people of lower socioeconomic status were at a higher risk of mental disorders and mood disorders [28]. Therefore, longer physical activity time, including work, may be associated with higher stress at work, leading to an increase in mentally unhealthy days.

As for the days with activity limitation, the results suggest that less than 60 minutes/day of exercise can reduce the number of activity limitation days. However, this relationship was different from that found for the other indicators of HRQOL (self-rated health, physically unhealthy days, mentally unhealthy days), where daily exercise, even for a long time, may have a positive impact. This difference may be because daily exercise for a long time increases the risk of injury that may interfere with daily life. Studies have shown that the average recreational runner suffers an injury rate of 2.5 to 12.1/1000 hours during running, which is one of the most popular forms of exercise [29]. In addition, workers who had no exercise habits and no physical activity time had more days with activity limitation, so it is especially important for workers who spend little time engaged in some form of physical activity to exercise during their leisure time.

These results indicate that it is crucial to have an exercise habit, regardless of the amount of time spent engaged in physical activity (including work), to have a good health-related quality of life. In addition, the results suggested that between 30 and 60 minutes of exercise daily was the most effective amount of exercise time for having a good health-related quality of life. Conversely, daily exercise, even for a short time, is thought to improve self-rated health and decrease mentally unhealthy days. Therefore, encouraging workers with no exercise habit to exercise daily even for a short time may benefit their health and work productivity.

### Limitations

This study has four limitations. First, the generalizability of the results is unclear because the CORoNaWork study is an Internet-based survey. However, to reduce sampling bias, sampling was conducted by generation, residence, and occupation. Second, because this was a cross-sectional study, the causal relationship between exercise time, physical activity time, and HRQOL could not be determined. Further research is needed to investigate the causal relationships using experimental designs with workers or employees as the sample population. For example, Emerson et al. [30] demonstrated that the effects of an exercise and nutrition workplace wellness program reduced stress and improved participants’ quality of life. Third, the questionnaire only asked about the duration of exercise in leisure time and physical activity, including work, not for exact exercise intensity and physical activity intensity.

Fourth, this study was conducted during the COVID-19 pandemic. As such, the impact of exercise and physical activity on HRQOL needs to be considered within the environmental context of the pandemic, which might differ under normal circumstances. Considering that the duration of physical activity was affected by the COVID-19 pandemic and that the participants were anxious about COVID-19 infection, a continuous evaluation is necessary.

## Conclusions

This study investigated the relationship between exercise time in leisure time, physical activity time, and HRQOL. These results suggest that daily exercise has a positive impact on HRQOL. Interventions to encourage workers with no exercise habit to exercise daily, even for a short time, may be an effective strategy to improve both workers’ health and work productivity during the COVID-19 pandemic.

## Data Availability

Due to the nature of this research, the participants of this study did not agree to their data being publicly shared, and hence, supporting data was not available.

## List of abbreviations

HRQOL: health-related quality of life
WHO: World Health Organization
COVID-19: coronavirus disease 2019
CDC: Centers for Disease Control and Prevention
LMM: linear mixed model
EMM: estimated marginal means

## Declarations

### Ethics approval and consent to participate

This study was approved by the Ethics Committee of the University of Occupational and Environmental Health, Japan (No. R2-079). Informed consent was obtained from the participants’ website.

### Consent for publication

Not applicable.

### Competing interests

The authors declare that they have no competing interests.

### Funding

This study was funded by a research grant from the University of Occupational and Environmental Health, Japan; a general incorporated foundation (Anshin Zaidan) for the development of educational materials on mental health measures for managers at small-sized enterprises; Health, Labour and Welfare Sciences Research Grants including Comprehensive Research for Women’s Healthcare (H30-josei-ippan-002) and Research for the Establishment of an Occupational Health System during a disaster (H30-roudou-ippan-007); and scholarship donations from Chugai Pharmaceutical Co., Ltd.

### Authors’ contributions

RS wrote the manuscript and analyzed the data. KI reviewed the manuscript, created the questionnaire, analyzed data, and provided advice on interpretation. HE,□MT,□ST,□TN,□and SM reviewed the manuscript. YF reviewed the manuscript and contributed to overall survey planning, creating the questionnaire, and securing funding for research. AO reviewed the manuscript, analyzed data, provided advice on interpretation, and secured funding for research.

## Acknowledgements

We appreciate all the participants and members of the CORoNaWork Study Group. The current members of the CORoNaWork Project, in alphabetical order, are as follows: Dr. Yoshihisa Fujino (present chairperson of the study group), Dr. Akira Ogami, Dr. Arisa Harada, Dr. Ayako Hino, Dr. Hajime Ando, Dr. Hisashi Eguchi, Dr. Kazunori Ikegami, Dr. Kei Tokutsu, Dr. Keiji Muramatsu, Dr. Koji Mori, Dr. Kosuke Mafune, Dr. Kyoko Kitagawa, Dr. Masako Nagata, Dr. Mayumi Tsuji, Ms. Ning Liu, Dr. Rie Tanaka, Dr. Ryutaro Matsugaki, Dr. Seiichiro Tateishi, Dr. Shinya Matsuda, Dr. Tomohiro Ishimaru, and Dr. Tomohisa Nagata. All members were affiliated with the University of Occupational and Environmental Health, Japan. We would also like to thank Editage (www.editage.com) for English language editing.

## Notes

### Competing Interest Statement

The authors have declared no competing interest.

## References

1. Bull FC, Al-Ansari SS, Biddle S, Borodulin K, Buman MP, Cardon G, Carty C, Chaput J-P, Chastin S, Chou R, et al. World Health Organization 2020 guidelines on physical activity and sedentary behaviour. British Journal of Sports Medicine.2020; 54:1451–1462.

2. Schuch FB, Vancampfort D, Richards J, Rosenbaum S, Ward PB, Stubbs B. Exercise as a treatment for depression: A meta-analysis adjusting for publication bias. Journal of Psychiatric Research. 2016;77:42–51.

3. Inoue M, Iso H, Yamamoto S, Kurahashi N, Iwasaki M, Sasazuki S, Tsugane S. Daily total physical activity level and premature death in men and women: results from a large-scale population-based cohort study in Japan (JPHC study). Ann Epidemiol. 2008;18:522–530.

4. Goodwin RD. Association between physical activity and mental disorders among adults in the United States. Prev Med. 2003;36:698–703.

5. Guthold R, Stevens GA, Riley LM, Bull FC. Worldwide trends in insufficient physical activity from 2001 to 2016: a pooled analysis of 358 population-based surveys with 1.9 million participants. The Lancet Global Health. 2018;6:e1077–e1086.

6. Ministry of Health Labour and Welfare Japan. Comprehensive Survey of Living Conditions (2018). https://www.mhlw.go.jp/content/10900000/000688863.pdf. Accesed 16 May 2021.

7. Matsuo T, So R. Socioeconomic status relates to exercise habits and cardiorespiratory fitness among workers in the Tokyo area. J Occup Health. 2021;63:e12187.

8. López-Bueno R, Smith L, Andersen LL, López-Sánchez GF, Casajús JA. Association between physical activity and sickness absenteeism in university workers. Occupational Medicine. 2019;70:24–30.

9. Walker TJ, Tullar JM, Diamond PM, Kohl HW 3rd, Amick BC 3rd. The Relation of Combined Aerobic and Muscle-Strengthening Physical Activities With Presenteeism. J Phys Act Health. 2017;14:893–8.

10. Walker TJ, Tullar JM, Diamond PM, Kohl HW, 3rd, Amick BC, 3rd. The longitudinal relation between self-reported physical activity and presenteeism. Prev Med. 2017;102:120–6.

11. Cindrich SL, Lansing JE, Brower CS, McDowell CP, Herring MP, Meyer JD. Associations Between Change in Outside Time Pre- and Post-COVID-19 Public Health Restrictions and Mental Health: Brief Research Report. Front Public Health. 2021;9:619129.

12. Balanzá-Martínez V, Kapczinski F, de Azevedo Cardoso T, Atienza-Carbonell B, Rosa AR, Mota JC, De Boni RB. The assessment of lifestyle changes during the COVID-19 pandemic using a multidimensional scale. Rev Psiquiatr Salud Ment. 2021;14:16–26.

13. Office for Novel Coronavirus Disease Control, Cabinet Secretariat, Government of Japan. COVID-19 Information and Resouces Declaration of State of Emergency. https://corona.go.jp/news/pdf/kinkyujitai_sengen_0407.pdf. Accesed 15 May 2021.

14. Centers for Disease Control and Prevention of USA. Health-Related Quality of Life (HRQOL) HRQOL Concepts. https://www.cdc.gov/hrqol/concept.htm. Accesed 5 May 2021.

15. Centers for Disease Control and Prevention of USA. Measuring healthy days: Population assessment of health-related quality of life. 2001. https://www.cdc.gov/hrqol/pdfs/mhd.pdf. Accesed 5 May 2021.

16. Chimed-Ochir O, Mine Y, Fujino Y. Pain, unhealthy days and poor perceived health among Japanese workers. J Occup Health. 2020;62:e12092.

17. Chimed-Ochir O, Mine Y, Okawara M, Ibayashi K, Miyake F, Fujino Y. Validation of the Japanese version of the CDC HRQOL-4 in workers. J Occup Health. 2020;62:e12152.

18. Fujino Y, Ishimaru T, Eguchi H, Tsuji M, Seichiro T, Ogami A, Mori K, Matsuda S. Protocol for a nationwide Internet-based health survey in workers during the COVID-19 pandemic in 2020. Journal of UOEH. 2021 (in print).

19. Centers for Disease Control and Prevention of USA. Health-Related Quality of Life (HRQOL) Methods and Measures. https://www.cdc.gov/hrqol/methods.htm. Accesed 5 May 2021.

20. Hsieh HH, Chang CM, Liu LW, Huang HC. The Relative Contribution of Dietary Habits, Leisure-Time Exercise, Exercise Attitude, and Body Mass Index to Self-Rated Health among College Students in Taiwan. Int J Environ Res Public Health. 2018; doi.org/10.3390/ijerph15050967.

21. Abuladze L, Kunder N, Lang K, Vaask S. Associations between self-rated health and health behaviour among older adults in Estonia: a cross-sectional analysis. BMJ Open. 2017;7:e013257.

22. Södergren M, Sundquist J, Johansson SE, Sundquist K: Physical activity, exercise and self-rated health: a population-based study from Sweden. BMC Public Health. 2008;8:352.

23. Brown DW, Brown DR, Heath GW, Balluz L, Giles WH, Ford ES, Mokdad AH. Associations between physical activity dose and health-related quality of life. Med Sci Sports Exerc. 2004;36:890–6.

24. Saint-Maurice PF, Troiano RP, Matthews CE, Kraus WE: Moderate-to-Vigorous Physical Activity and All-Cause Mortality: Do Bouts Matter? J Am Heart Assoc. 2018; doi: 10.1161/JAHA.117.007678.

25. Wilson JJ, Blackburn NE, O’Reilly R, Kee F, Caserotti P, Tully MA. Association of objective sedentary behaviour and self-rated health in English older adults. BMC Research Notes. 2019; doi: 10.1186/s13104-019-4050-5.

26. Munehiro M, Masaki T, Takashi A. Association of work and travel-related and recreational physical activity with depression among Japanese adults: cross-sectional study. JSLS. 2016;13:35–41.

27. Beenackers MA, Kamphuis CBM, Giskes K, Brug J, Kunst AE, Burdorf A, van Lenthe FJ. Socioeconomic inequalities in occupational, leisure-time, and transport related physical activity among European adults: A systematic review. International Journal of Behavioral Nutrition and Physical Activity. 2012; doi: 10.1186/1479-5868-9-116.

28. Kivimäki M, Batty GD, Pentti J, Shipley MJ, Sipilä PN, Nyberg ST, Suominen SB, Oksanen T, Stenholm S, Virtanen M, et al. Association between socioeconomic status and the development of mental and physical health conditions in adulthood: a multi-cohort study. Lancet Public Health. 2020;5:e140–e149.

29. van Mechelen W. Running injuries. A review of the epidemiological literature. Sports Med. 1992;14:320–335.

30. Emerson ND, Merrill DA, Shedd K, Bilder RM, Siddarth P. Effects of an employee exercise programme on mental health. Occupational Medicine. 2016;67:128–134.

